# Thalamic stereoelectroencephalography: safety, accuracy, and thalamocortical connectivity

**DOI:** 10.64898/2025.12.01.25341375

**Authors:** Sihyeong Park, Gloria Ortiz-Guerrero, Fiona Permezel, Hicham Dabaja, Gamaleldin Osman, Brian Burkett, Steven Messina, Keith Starnes, Brian N. Lundstrom, Lily C. Wong-Kisiel, David Burkholder, Dora Hermes, Benjamin H. Brinkmann, Gregory Worrell, W. Richard Marsh, Kai J. Miller, Jamie J. Van Gompel, Nicholas M. Gregg

**Author notes:** Correspondence to: Nicholas Gregg, Sihyeong Park, Department of Neurology, Mayo Clinic, 200 First Street SW. Rochester, MN.

## Abstract

**Background and Objectives:** There is growing interest in thalamic sampling during epilepsy stereoelectroencephalography (sEEG). Data on thalamic sEEG safety, accuracy, and anterior (ANT), centromedian (CM), and pulvinar (PUL) nucleus connectomics is scarce. Here, we report the safety, accuracy and connectivity of thalamic sEEG and compare results to epilepsy DBS.

**Methods:** This single-center study included all patients with epilepsy implanted with thalamic sEEG or DBS between 2018 and 2024. Stereo-EEG surgery used a frameless stereotactic articulating arm, 3D printed stereotactic guide, or robot (sEEG lead diameter 0.8mm). DBS surgery used a rigid frame and cannula (1.81mm outer diameter). Post-surgical complications were identified by chart and imaging review. For each thalamic lead, accuracy was defined as the distance from the nearest electrode contact to the targeted nucleus Morel atlas volume (ANT, CM, or PUL), using an open-source toolbox. Estimated volumes of tissue activation/sampling were calculated for each most proximate contact for thalamocortical connectivity analysis. Connectivity analyses used a normative structural connectome, and connectivity patterns were compared.

**Results:** 160 thalamic sEEG leads and 188 DBS leads were implanted across 109 and 83 patients, respectively. One sEEG patient (0.9%) developed a symptomatic intraparenchymal hemorrhage with transient weakness of the contralateral upper extremity. Eight patients with DBS had transient post-surgical symptomatic complications, three with radiographic findings. Targeting accuracy was excellent for sEEG and DBS with median proximity of 0.30mm and 0.23mm, respectively; sEEG was associated with greater variability and more outliers (proximity≥3mm; 4% vs. 0, *p*=0.004). Thalamocortical connectivity patterns were highly consistent between sEEG and DBS cohorts with excellent overlap for ANT, CM and PUL subgroups (Spearman’s ρ=0.86-0.98, *p*<1e^-5^). The ANT subgroup showed preferential connectivity to prefrontal and mesial temporal cortices; CM with perirolandic cortex, supplementary motor area, and subcortical regions; and PUL with mesial and neocortical temporal, and parieto-occipital regions, with some territories of overlap or under-engagement.

**Discussion:** Thalamic sEEG demonstrates a favorable safety profile, excellent targeting accuracy, and representative thalamocortical network engagement relative to DBS. Favorable sEEG safety profile may reflect difference in lead cross-sectional area (5.1-fold smaller than DBS cannula). Distinct ANT, CM, and PUL thalamocortical connectivity profiles support individualized, hypothesis-driven targeting.

## Introduction

Stereoelectroencephalography (sEEG) is a crucial tool for evaluating seizure networks in patients with drug-resistant focal epilepsy. Multi-contact sEEG leads are implanted in a hypothesis-driven manner to delineate pathological and functional networks. Stereo-EEG applications include identification of seizure onset regions and propagation patterns, stimulation mapping to assess eloquent cortex and for seizure provocation, and effective connectivity mapping.^1^

Traditionally, sEEG investigations have focused on cortical structures. However, growing evidence highlights the thalamus as a critical node in seizure initiation and propagation.^2,3^ Nucleus-specific connectivity profiles,^4–6^ the prognostic value of thalamic epileptogenicity,^7^ and the rise of network-guided neuromodulation with non-standard stimulation targets^5^ has driven increasing interest in single- and multi-site thalamic sEEG. Thalamic sEEG applications include characterization of subcortical seizure network nodes,^7^ seizure lateralization,^8^ identification of ictal thalamic signatures to guide responsive neurostimulation (RNS) and deep brain stimulation (DBS) detectors,^9–11^ mapping thalamo-cortical effective connectivity with single-pulse stimulation,^12–14^ and thalamic stimulation trials aimed at informing neuromodulatory device^15^ targeting, programming, adaptive paradigms, and connectomic DBS approaches.^16^

While previous work has investigated the electrophysiology of the thalamus under clinical and research indications, aiming to understand thalamic activity during seizures,^17^ data on thalamic sEEG safety and accuracy is scarce with limited numbers of patients. In one series of 24 patients implanted in the anterior, mediodorsal, or centromedian nuclei, asymptomatic hemorrhage or edema was noted in 34%.^18^ Another study^4^ reported on multisite thalamic sEEG feasibility and safety (*n*=11) with no complications, while a recent retrospective cohort study showed no statistical difference in complication rates between thalamic (*n*=30) and non-thalamic sEEG groups.^19^

Here, we report on the safety and targeting accuracy of thalamic sEEG in 109 patients (160 leads), with comparison 83 patients (188 leads) undergoing DBS for epilepsy, and delineate thalamocortical network engagement across anterior (ANT), centromedian (CM), and pulvinar (PUL) nuclei to inform network-guided thalamic investigations.

## Methods

### Study Design

This single center, observational retrospective cohort study included all patients with drug-resistant epilepsy implanted with thalamic sEEG or DBS for drug resistant epilepsy between 2018 and 2024. This study was approved by the Mayo Clinic Institutional Review Board. The initial thalamic sEEG practice at our institution was performed under an IRB approved protocol in which existing clinical leads were extended into the thalamus (*n*=11). As these leads did not have thalamic nucleus-specific targeting, these patients were excluded from this analysis.

The remaining patients had thalamic leads placed as part of clinical practice, with hypothesis-driven thalamic nucleus selection (generally, anterior nucleus of thalamus (ANT) for frontotemporal seizure networks; centromedian nucleus (CM) for peri-rolandic or broad/bihemispheric seizure networks; or pulvinar nucleus (PUL) for posterior quadrant or neocortical temporal seizure networks), decided at a multidisciplinary surgical epilepsy conference. Thalamic leads are often considered by our clinical team for patients with suspected poorly localized, highly eloquent, or multifocal seizure networks, when chronic treatment with a neuromodulation therapy is likely. Six patients were excluded due to significant structural brain abnormalities that prevented reliable image post-processing for thalamic atlas normalization.

### Surgical Method

sEEG lead placement used a Medtronic precision aiming device frameless stereotactic articulating arm, Starfix 3D printed stereotactic guide, or Renishaw robotic system; DBS surgery was performed using the Leksell frame and rigid cannula (Figure 2B). Intermittent robotic system use started in 2022.

### Demographics, Complications

Demographics, seizure outcomes, and acute symptomatic surgical complications were obtained by chart review. Postoperative CT reports were reviewed to identify radiographic findings (including asymptomatic radiographic findings, and symptomatic imaging-positive complications). For patients with radiographic findings, CT images were reviewed to identify the culprit electrodes.

### Image Processing, Lead Localization, Lead Accuracy

Imaging coregistration, normalization into Montreal Neurological Institute (MNI) space, and lead localization was performed using Lead-DBS version 3.2.^20^ Thalamic nuclei were defined by the Morel atlas.^21^ Targeting accuracy was assessed for a single electrode contact from each thalamic lead. Accuracy was defined as the Euclidean distance from the nearest electrode contact to the Morel atlas representation of the targeted nucleus (ANT, CM, or PUL). The ANT included both the anteromedial and anteroventral nuclei. Reported distances are measured from the center of the nearest electrode contact to the closest vertex on or within the 3D surface-based volume of the target nucleus.

### Statistics

Boxplots illustrate the proximity values in each cohort. Distances <1.5 mm were considered optimal; outliers were defined as contact-to-target proximity values ≥3.0 mm.^22^ Wilcoxon rank-sum test assessed group level comparison of proximity values. To assess whether targeting accuracy was affected by the implant complexity, electrodes were stratified by the number of thalamic targets/leads per case, and proximity values were compared using Kruskal-Wallis test. Kruskal-Wallis test compared the accuracy between surgical techniques. Pairwise comparisons were performed using Wilcoxon rank-sum test with Bonferroni correction for multiple comparisons. For subgroup analysis, patients were stratified into pediatric (<18 years) and adult (≥18 years) subgroups, and proximity values were compared using the Wilcoxon rank-sum test. To assess accuracy across time, linear regression was performed with implant year as predictor variable. Levene’s test assessed the equality of variances of the proximity values in sEEG and DBS cohorts. For comparison of the outlier burden between the surgical techniques, Pairwise Fisher test was performed with Bonferroni correction. Statistical analysis was performed using MATLAB 2023a.

### Connectivity Analysis

For each sEEG and DBS thalamic lead, the electrode contact that was closest to the target was used for connectivity analysis, using Lead-DBS version 3.2. A region of interest (ROI) was generated for each contact, which was defined as the modeled volume of tissue activated (VTA) by a 3.0 mA stimulus^23^ (this was used for patient-specific connectivity analysis demonstrated in Figure 3H and 5). This approach should better reflect the sampled tissue when compared to a model based on uniform contribution from the geometric center of the contact. Calculated regions for each electrode were taken to calculate a weighted center of mass (COM) for each target and electrode type

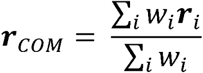

where ***r****_COM_* is the coordinate vector [*x_COM_*, *y_COM_*, *z_COM_*]*^T^* of the center of mass, ***r****_i_* are the coordinates [*x_i_*, *y_i_*, *z_i_*]*^T^* of each voxel that constitute the volumes of tissue engagement, and ***w****_i_* are the weights derived from the intensity of activation of each voxel. A center of mass ROI was defined as a 2 mm radius sphere around the weighted center of mass (this was used for group-level fiber filtering illustrated in Figure 3A-G), based on previous work on subcortical local field potentials.^24^ Fibers were seeded from each center of mass ROI using a structural normative connectome of healthy subjects.^25^ Visualizations used a 7 Tesla MRI ex vivo human brain template^26^ and Surf Ice surface rendering tool (https://www.nitrc.org/projects/surfice/).

For quantitative analysis of the thalamocortical fibers, the Klein cortical parcellation atlas was used,^27^ which contains 31 cortical parcels in each hemisphere. As the Klein atlas does not include the hippocampus or amygdala, these two parcels were taken from the Automated Anatomical Labeling atlas 3 (AAL3).^28^ Since VTAs were mirrored to the left hemisphere for connectivity analysis, only left-sided parcels from this composite parcellation atlas were used.

Then, connectivity was assessed between each patient’s thalamic ROI and the composite Klein/AAL3 atlas, using the normative structural connectome.^25^ Connectivity matrices were then used for group level comparison of the thalamocortical connectivity patterns between sEEG and

DBS cohorts. Wilcoxon rank-sum was used for group level comparison of the parcel-specific connectivity. *p*-values were adjusted using Benjamini Hochberg false discovery rate (FDR) correction. Statistical significance was defined as FDR-adjusted *p* (*p_FDR_*) < 0.05. Spearman’s rank correlation assessed the similarity of the parcel connectivity ranks between the sEEG and DBS cohorts.

### Data Availability

Data will be made available upon reasonable request.

## Results

### Patient Demographics

A total of 160 thalamic sEEG leads were implanted (124 ANT, 27 CM, 9 PUL) in 109 patients (53% female) (Table 1), 56 (51.4%) of whom were subsequently recommended for intracranial neuromodulation at the post-sEEG multidisciplinary surgical epilepsy consensus conference. A total of 188 DBS electrodes were implanted (140 ANT, 35 CM, 13 PUL) in 83 patients (63.9% female) (including 2- and 4-lead DBS systems). Median age at implantation was 23 and 27 years for the sEEG and DBS cohorts, respectively.

**Table 1.** Patient demographics and accuracy.

| Characteristic | sEEG total | sEEG ANT | sEEG CM | sEEG PUL | DBS total | DBS ANT | DBS CM | DBS PUL |
| --- | --- | --- | --- | --- | --- | --- | --- | --- |
| N, patients | 109 | 101 | 20 | 9 | 83 | 75 | 21 | 7 |
| Age >=18 | 78 | 72 | 9 | 7 | 67 | 62 | 11 | 6 |
| Age <18 | 31 | 29 | 10 | 2 | 16 | 13 | 10 | 1 |
| N, electrodes | 160 | 124 | 27 | 9 | 188 | 140 | 35 | 13 |
| Age at implant,<br>(years),<br>*median [IQR] | 23<br>[17, 35] | 23<br>[17, 35] | 16.5<br>[14, 25] | 27<br>[18, 37] | 27<br>[18, 41] | 27<br>[18,41] | 18<br>[15,28] | 30<br>[24.5,39] |
| Age >=18 | 26.5<br>[23, 38] | 26<br>[22, 37] | 25<br>[23, 37] | 33<br>[25,38.5] | 31.5<br>[25,43] | 30<br>[24,43] | 28<br>[24,36.5] | 32.5<br>[26,41] |
| Age <18 | 15<br>[12, 16] | 15<br>[12, 16] | 15<br>[12.5,16] | - | 16.0<br>[15,18] | 15<br>[15,16] | 15<br>[12,16] | - |
| Proximity<br>(mm),<br>*median [IQR] | 0.30<br>[0.21, 0.77] | 0.32<br>[0.24,1.06] | 0.26<br>[0.20,0.40] | 0.15<br>[0.12,0.18] | 0.23<br>[0.17,0.31] | 0.24<br>[0.17,0.33] | 0.22<br>[0.16,0.28] | 0.21<br>[0.16,0.23] |
| Age >=18 | 0.29<br>[0.21, 0.55] | 0.35<br>[0.25,1.13] | 0.31<br>[0.21,0.50] | 0.17<br>[0.15,0.19] | 0.23<br>[0.17,0.35] | 0.24<br>[0.17,0.33] | 0.23<br>[0.19,0.28] | 0.21<br>[0.17,0.24] |
| Age <18 | 0.26<br>[0.20, 0.42] | 0.28<br>[0.19,0.49] | 0.22<br>[0.18,0.33] | - | 0.22<br>[0.15,0.26] | 0.24<br>[0.18,0.33] | 0.18<br>[0.16,0.24] | - |

\*Median [1<sup>st</sup> quartile, 3<sup>rd</sup> quartile].

### Comparison of Thalamic sEEG and DBS Complications

Transient symptomatic imaging-positive complications occurred in both cohorts. One sEEG patient (0.9%) developed a symptomatic intraparenchymal hemorrhage in the posterior limb of the right internal capsule, with transient left arm weakness, with recovery to baseline prior to discharge. Three patients with DBS (3.7%) had symptomatic imaging-positive complications, all of which were transient: one patient (1.2%) had tract edema and seizures of new semiology, which resolved prior to discharge; two patients (2.4%) had small intraparenchymal hemorrhage involving the internal capsule, with mild contralateral weakness, which returned to baseline.

Five DBS patients had acute post-surgical complications without postoperative imaging findings. One patient with ANT-DBS developed post-implantation psychosis prior to activation of stimulation, which required hospital evaluation and resolved spontaneously over two days. One ANT-DBS patient had worsening of preexisting upper motor neuron pattern facial weakness, which was treated with steroids and returned to pre-operative baseline. Three DBS patients (two ANT-DBS, one CM-DBS) had a transient increase in seizure frequency.

Asymptomatic radiographic findings were observed in each cohort. In the sEEG cohort, six patients (5.4%) had mild postoperative intracranial hemorrhage (one epidural hemorrhage, four subarachnoid hemorrhage, and one intraventricular hemorrhage). The thalamic lead was implicated in the intraventricular hemorrhage. A left parietal subarachnoid hemorrhage occurred in one patient; a pulvinar lead and two non-thalamic leads traversed the region, with possible attribution to the thalamic lead. Intracranial hemorrhage in the other four patients was not associated with thalamic sEEG leads.

In the DBS cohort, three patients (3.6%) had asymptomatic trace intracranial hemorrhage after implantation (two intraparenchymal hemorrhage, one subarachnoid hemorrhage), all attributed to DBS lead placement.

### Targeting Accuracy of Thalamic sEEG and DBS

Figure 1 shows group renderings of thalamic sEEG and DBS lead locations for each target nucleus. Median proximity of sEEG electrodes was 0.30 mm [IQR=0.21-0.77], indicating that most leads had a contact within or in close proximity to the atlas volume of the targeted nucleus. Target-specific median sEEG contact proximity to ANT, CM, PUL was 0.32 mm [IQR=0.24-0.94], 0.26 mm [IQR=0.20-0.40], and 0.15 mm [IQR=0.12-0.18], respectively. In the DBS cohort, median proximity was 0.23 mm [IQR=0.17-0.30]. Target-specific median proximity to ANT, CM, PUL was 0.24 mm [IQR=0.17-0.33], 0.22 mm [IQR=0.16-0.28], and 0.21 mm [IQR=0.16-0.23], respectively.

**Figure 1.**
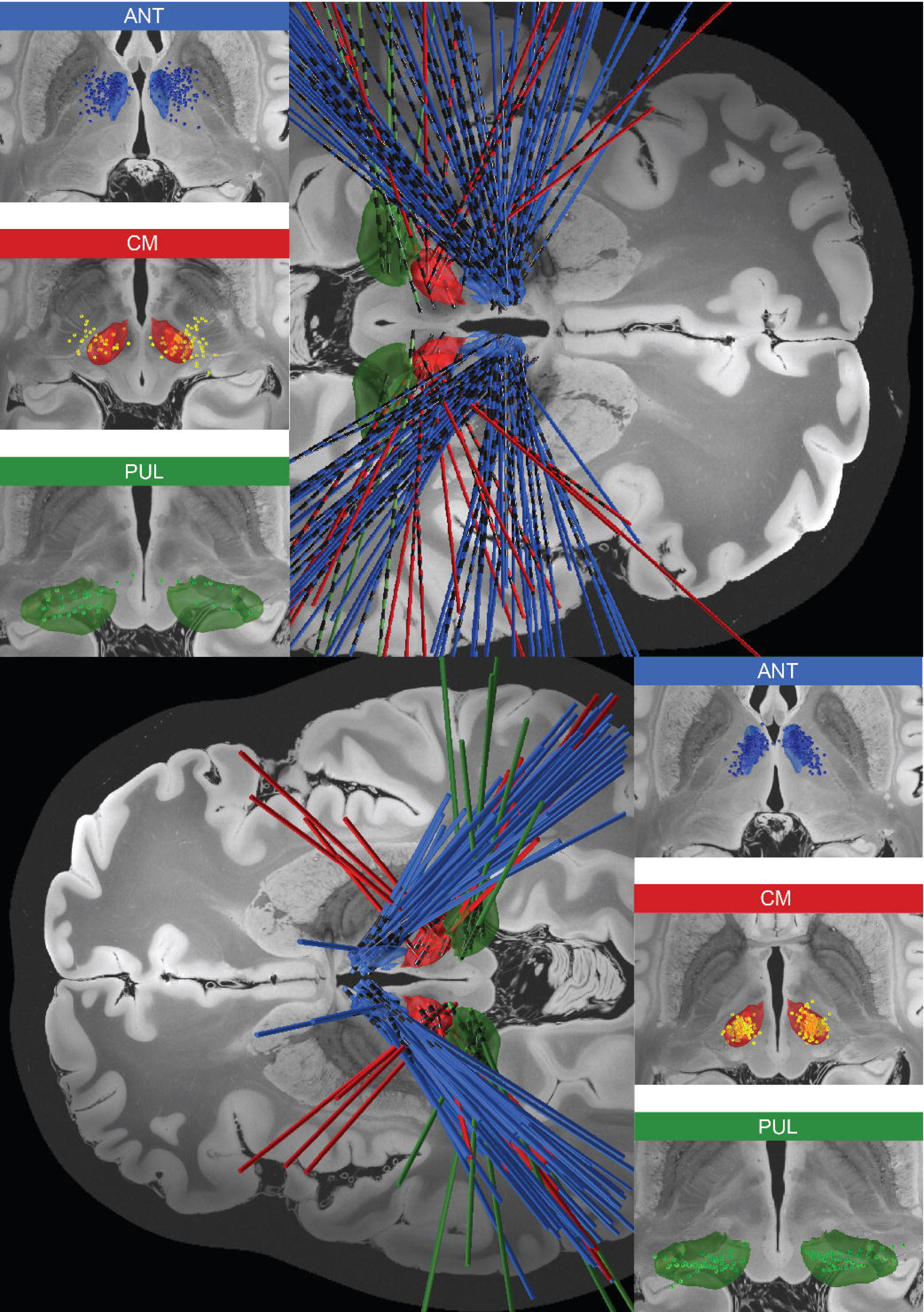
Group renderings of the sEEG electrodes and their contacts implanted in three different thalamic nuclei. Results are visualized in 7 Tesla MRI of the ex vivo human brain template^26^ and Morel atlas.^21^ **Top panel**: Left upper and middle panel: three deepest contacts of sEEG electrodes in blue (ANT) and yellow (CM). Left lower panel: four deepest contacts of sEEG electrodes in green (PUL). Note longer contact length and spacing for the sEEG electrodes (2 mm, 1.5 mm) vs DBS electrodes (1.5 mm, 0.5 mm). Right panel: Full trajectories of 160 sEEG electrodes implanted in 109 patients. **Bottom panel**: Right panel: group renderings of the DBS electrodes and their contacts implanted in ANT (blue), CM (red), and PUL (green). Left panel: full trajectories of 187 DBS electrodes implanted in 82 patients. One subject had a CM-DBS electrode with cross-hemisphere trajectory due to lack of ipsilateral bone anchor from previous brain surgery. This patient was withheld from visualization to avoid confusion; lead shown in Figure S4.

The sEEG and DBS contact-target proximities were comparable for the CM (*p*=0.059) and PUL cohorts. All PUL sEEG and DBS leads had contacts within the nucleus; the difference in calculated proximities (0.06 mm greater for DBS than sEEG leads; *p*=0.038) is due to the larger diameter of the DBS lead vs. sEEG lead (1.36 mm vs. 0.8 mm) and the analysis method that measures proximity from contact center to closest atlas vertex (Figure 2A). ANT targeting did show greater contact-target separation in the sEEG vs DBS cohort (0.08 mm; *p*=7e-06).

**Figure 2.**
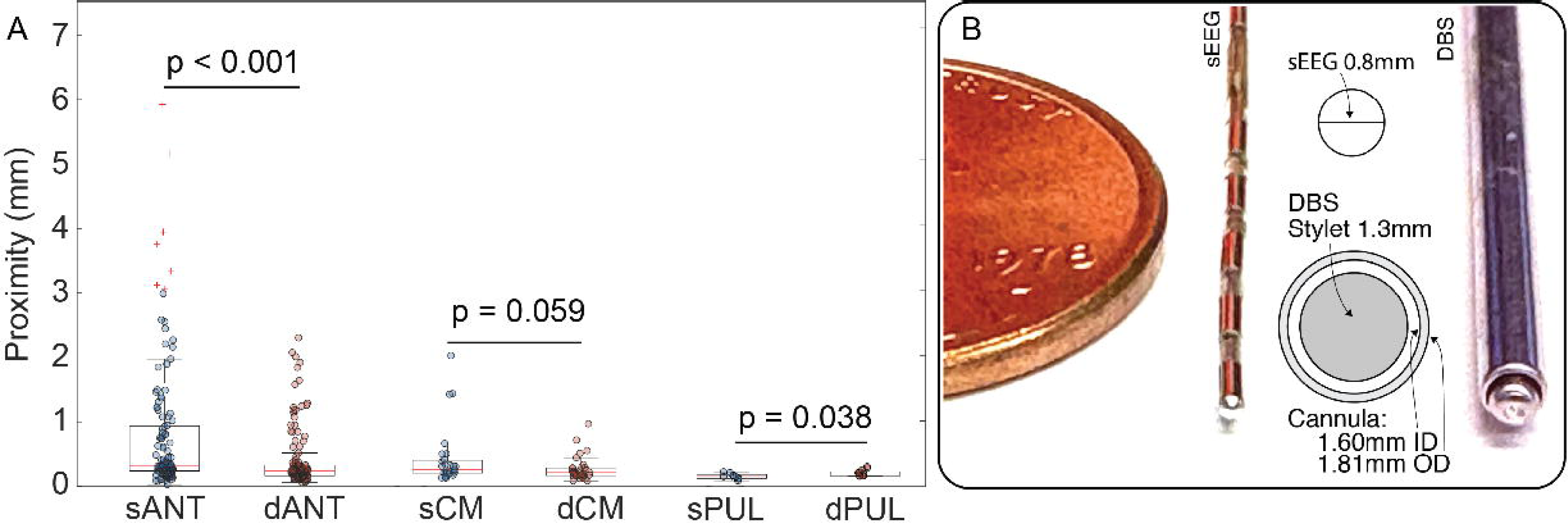
**(A) Boxplots and scatter plots of group level comparison of thalamic sEEG and DBS accuracy.** sANT, dANT: ANT subgroup in sEEG and DBS cohorts. sCM, dCM: CM subgroup in sEEG and DBS cohorts. sPUL, dPUL in sEEG and DBS cohorts. **(B) Comparison of sEEG electrode and DBS surgery stylet and rigid cannula system.** sEEG electrode and DBS surgery cannula/stylet system are shown, to scale, next to a penny. A commonly used sEEG electrode has 0.8mm diameter, with corresponding cross-sectional area of 2.01 mm^2^. A standard cannula/stylet system for DBS surgery with cannula outer diameter of 1.81 mm (cross-section area: 10.29 mm^2^; 5.1-fold larger area vs. sEEG).

Overall, there were more targeting outliers in the sEEG cohort compared to the DBS cohort (≥3 mm; 7/160 vs 0/188, Fisher test *p*=0.004), and more sEEG contacts than DBS contacts fell outside of optimal targeting accuracy (≥1.5 mm; 19/160 vs 7/188, *p*=0.006). This is reflected in greater variability in contact-target proximities in the sEEG cohort vs DBS (Levene’s test, *p*=7e-11), which was largely driven by the ANT subgroup. ANT sEEG had seven outliers (defined as proximity value ≥3 mm) vs. no outliers in the ANT DBS subgroup. There were no outliers in the CM or PUL subgroups.

### Thalamic sEEG Accuracy Subgroup Analyses – Surgical Technique, Targeting (Single vs Multi-target), and Age (Pediatric vs Adult)

The accuracy of sEEG by surgical implantation technique is shown in Figure 4. Median proximity values for the stereotactic articulating arm, 3D printed stereotactic guide, or robotic system were 0.34 mm [IQR=0.23-1.04] (*n*=54), 0.32 mm [IQR=0.22-1.20] (*n*=22), 0.28 mm [IQR=0.19-0.41] (*n*=84), respectively (*p*=0.09), with 5 outliers in the articulating arm subgroup (9.3%), and 1 outlier each in the 3D printed guide (4.6%) and robotic system (1.2%) subgroups. Single vs multitarget thalamic sEEG accuracy was not significantly different (*p*=0.25), with 71 leads placed in single thalamic target cases, 64 leads in 2-target cases, and 27 leads in 4-target cases. Targeting accuracy for pediatric and adult patients was comparable, across 109 leads implanted in 80 adults and 53 leads in 31 pediatric patients, with median proximity of 0.31 mm [IQR=0.20-1.00] and 0.26 mm [IQR=0.20-0.42], respectively (*p*=0.065). Detailed subgroup analyses are reported in Supplementary Materials.

### Thalamocortical Connectivity in sEEG and DBS

The sEEG and DBS group-level weighted center of mass ROIs are shown in Figure 3A-C for ANT, CM, and PUL subgroups. Group-level weighted center of mass ROI-filtered thalamocortical connectivity was visualized for sEEG (Figure 3) and DBS (Figure S1 and Figure S2) cohorts. Euclidian distance between the center coordinate of the sEEG center of mass and DBS center of mass for ANT, CM, and PUL were 1.1, 2.1, and 3.4 mm, respectively. Distinct thalamocortical connectivity profiles were observed between ANT, CM, and PUL subgroups.

**Figure 3.**
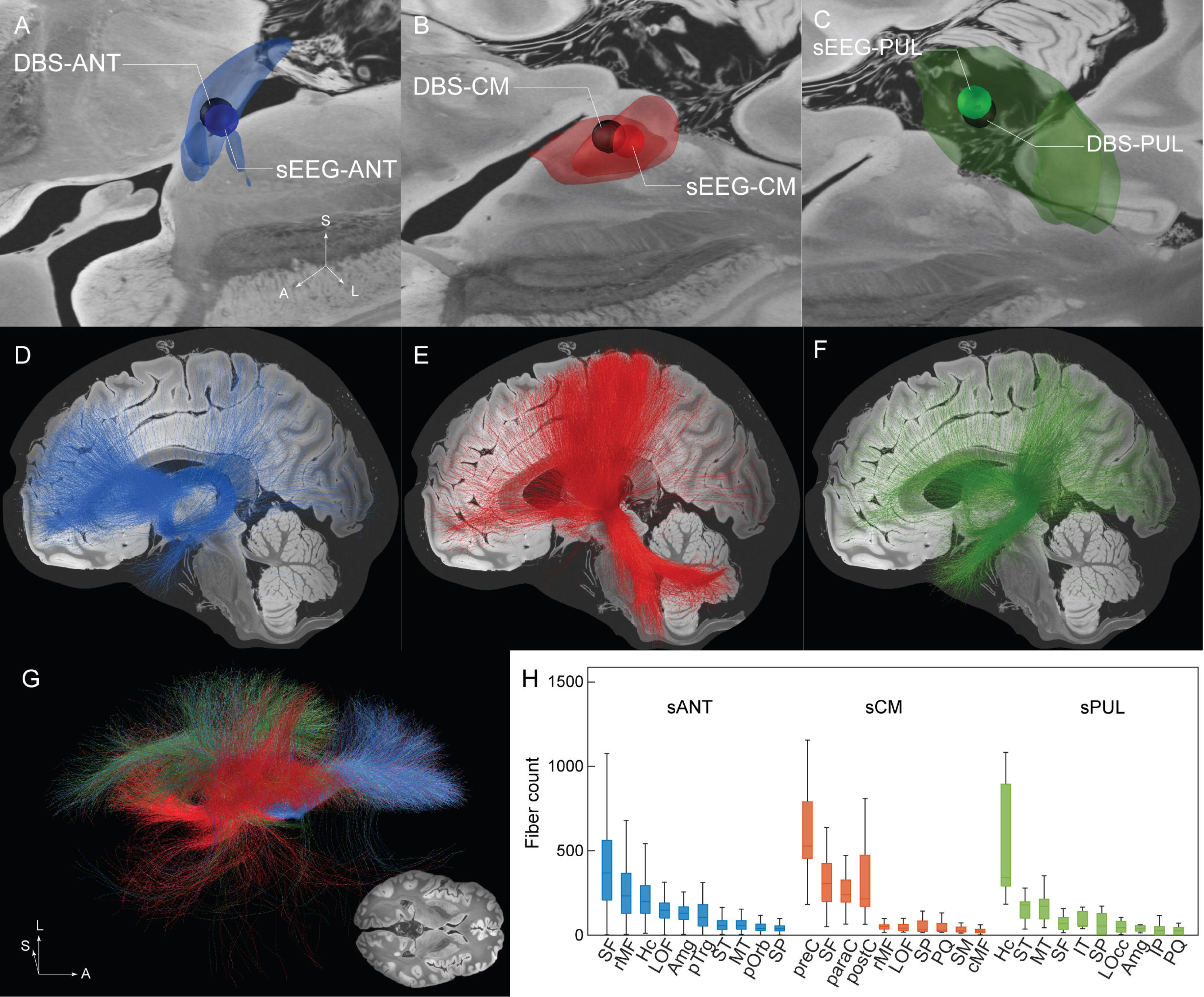
Visualization of the region of interest (ROI) created from the weighted center of mass (COM). (A) ROI of sEEG-ANT subgroup (blue) and DBS-ANT subgroup (black). (B) ROI of sEEG-CM subgroup (red) and DBS-CM subgroup (black). (C) ROI of sEEG-PUL subgroup (green) and DBS-PUL subgroup (black). (D-F) Thalamocortical structural connectivity seeded from the thalamic ROI for ANT-sEEG, CM-sEEG, PUL-sEEG. (G) Top-down view of thalamocortical connectivity from ANT (blue), CM (red), PUL (green) in sEEG cohort. (H) Thalamic target-specific connectivity profiles. Top 10 cortical parcels with highest median fiber count were plotted for each thalamic target. Amg: amygdala, cMF: caudal middle frontal, Hc: hippocampus, IP: inferior parietal, IT: inferior temporal, LOc: lateral occipital, LOF: lateral orbitofrontal, MT: middle temporal, paraC: paracentral, postC: postcentral, preC: precentral, PQ: precuneus, pOrb: pars orbicularis, pTrg: pars triangularis, rMF: rostral middle frontal, SF: superior frontal, SM: supramarginal, SP: superior parietal, ST: superior temporal.

**Figure 4.**
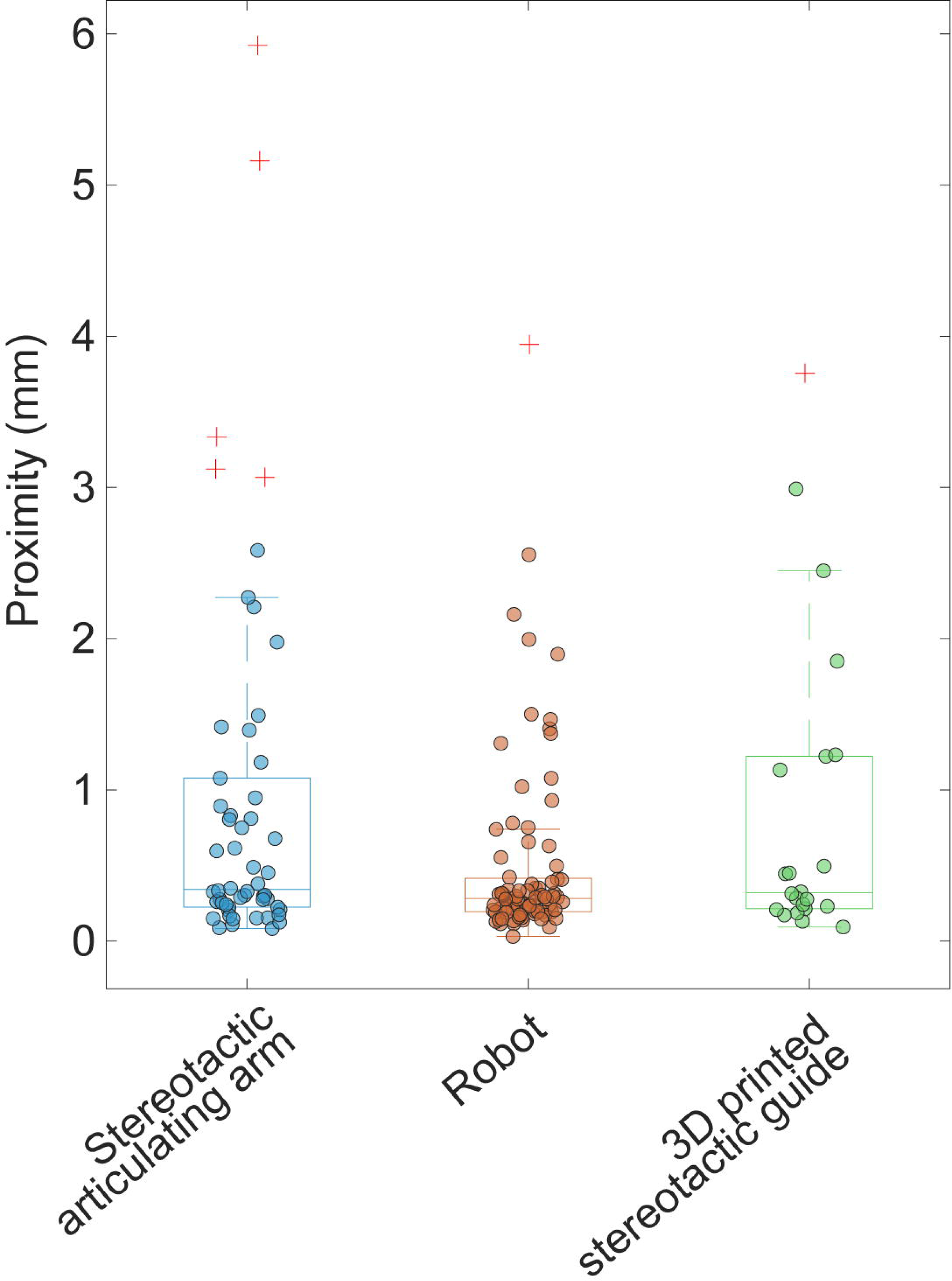
Comparison of accuracy of surgical techniques. Median proximity values for the stereotactic articulating arm, 3D printed stereotactic guide, or robotic system were 0.34 mm (*n*=54), 0.32 mm (*n*=22), 0.28 mm (*n*=84), respectively (*p*=0.09), with 5 outliers in the articulating arm subgroup (9.3%), and 1 outlier each in the 3D printed guide (4.6%) and robotic system (1.2%) subgroups.

For individual-level quantitative connectivity analysis, patient-specific VTAs were used. Thalamocortical fiber counts seeded from patient-specific thalamic ROIs were quantified for each parcel of the composite Klein + AAL3 amygdalohippocampal atlas. The fiber counts between the thalamic ROIs and each of the top 10 engaged parcels are listed in Figure 3H. ANT sEEG connectivity showed high fiber counts to prefrontal cortex and mesial temporal structures. CM sEEG showed preferential connectivity to peri-rolandic parcels, supplemental motor area, and superior frontal gyrus. PUL sEEG showed preferential connectivity to mesial and neocortical temporal cortices and parieto-occipital regions. Some overlapping connectivity was present; ANT and CM showed strong superior frontal gyrus connectivity with a rostral (favor ANT) caudal (favor CM) gradient. ANT showed stronger connectivity to the rostral middle frontal and lateral orbitofrontal cortices than CM. CM and PUL subgroups showed some overlapping projections to the parietal region; CM parietal connectivity was largely restricted to the postcentral gyrus. Visual comparison of the fibers between the sEEG and DBS subgroup for each thalamic target showed similar thalamocortical connectivity patterns (Figure S1).

Subject-level connectivity profiles were compared between the sEEG and DBS cohorts for the 10 parcels with the highest connectivity for each target subgroup (Figure 5). The relative strength and hierarchy of thalamocortical projections, assessed by Spearman’s ρ correlation of ranks, were well conserved for all subgroups: ANT ρ=0.98; CM ρ=0.98; and PUL ρ=0.86; all *p*<1.5e^-7^ (Figure 5). Thalamocortical connectivity between sEEG and DBS for the top 10 parcels did show absolute differences in fiber counts for ANT (all parcels) and CM (peri-rolandic parcels but not frontal parcels) subgroups. The PUL subgroup had no significant difference for any parcels (*p_FDR_* >0.05).

**Figure 5.**
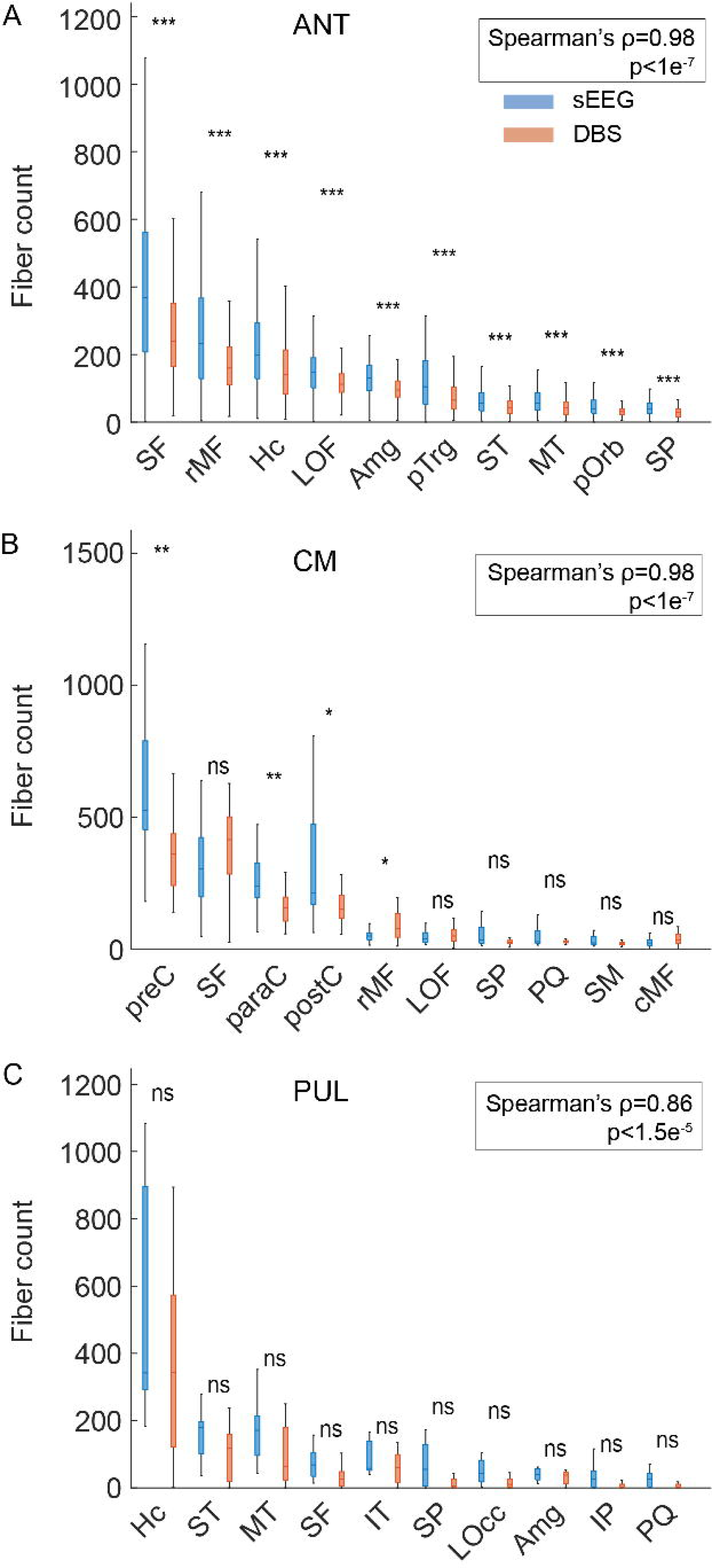
Group level comparison of thalamocortical connectivity. Non-parametric test was performed to compare the thalamocortical connectivity in sEEG (blue) and DBS (red) cohort in each parcel (A: ANT, B: CM, C: PUL). \**p* <0.05; \*\**p* <0.01; \*\*\**p* <0.001; ns, not significant. Top 10 cortical parcels with highest median fiber count were plotted for each thalamic target. Spearman’s ρ correlation of ranks tests the monotonic agreement in the order of thalamocortical connectivity between sEEG and DBS. Amg: amygdala, cMF: caudal middle frontal, Hc: hippocampus, IP: inferior parietal, IT: inferior temporal, LOc: lateral occipital, LOF: lateral orbitofrontal, MT: middle temporal, paraC: paracentral, postC: postcentral, preC: precentral, PQ: precuneus, pOrb: pars orbicularis, pTrg: pars triangularis, rMF: rostral middle frontal, SF: superior frontal, SM: supramarginal, SP: superior parietal, ST: superior temporal.

## Discussion

In this retrospective cohort study, we demonstrate that thalamic sEEG is associated with fewer symptomatic acute surgical complications compared to DBS for epilepsy, achieves high targeting accuracy, and reliably engages distinct large-scale brain networks across ANT, CM, and PUL nuclei. To our knowledge, this is the largest study to date to evaluate the safety and accuracy of thalamic sEEG, and provides important evidence to inform network-guided thalamic investigations.^5^ These findings are particularly timely, given the growing interest in network-guided neuromodulation and the current paucity of data on the safety, accuracy, and connectomics of thalamic sEEG and DBS for epilepsy.

Thalamic neuromodulation has entered a period of accelerated growth (51% of patients in the present sEEG cohort recommended for intracranial neuromodulation at surgical epilepsy conferences), and traditional DBS of the ANT for drug-resistant focal (limbic connected)^29^ epilepsy is increasingly complemented by alternative off-label subcortical targets.^30–38^ This is accompanied by an emerging framework of network-guided neuromodulation^5,39^ for epilepsy, and similarly connectomic DBS for movement disorders DBS.^40–43^ Our results demonstrate that thalamic sEEG connectivity is broadly representative of DBS, with rare exceptions, and provides detailed evidence on the safety and accuracy of thalamic sEEG, thereby supporting the feasibility of sEEG to explore thalamocortical networks and to derive patient-specific insights for chronic thalamic neuromodulation.

Regarding safety, the hemorrhagic complication rate in our sEEG cohort was comparable to meta-analysis estimates of non-thalamic sEEG of 1.0%,^44^ and consistent with prior thalamic series, with no thalamic hemorrhage observed.^4,18^ One transient intraparenchymal hemorrhage occurred along a thalamic trajectory involving the internal capsule, with complete resolution of contralateral upper extremity weakness prior to discharge from the epilepsy monitoring unit. Entry point and trajectory selection remain key determinants of risk. Additionally, surgical complications in the DBS cohort aligned with prior large series (e.g. 8.4% neurological complications, 2.8% symptomatic hemorrhage in 800 subthalamic DBS patients).^45^ The greater complication rate in DBS, despite superior accuracy, likely reflects instrumentation—specifically, the much larger (5.1-fold) cross-sectional area of DBS guide cannulae compared to sEEG leads, which are placed using an internal rigid stylet for trajectory control. Once seated, the stylet is withdrawn to leave a flexible implanted lead) (Figure 2B).

While both thalamic sEEG and DBS demonstrated high targeting accuracy across all nuclei, the sEEG cohort had greater variability, with more sub-optimal (electrode-target proximity ≥1.5mm) and outlier (≥3mm) leads. This likely reflects technical differences, as noted above: for DBS, the guide cannula creates a rigid track for lead insertion, whereas sEEG relies on an internal stylet to provide the stiffness needed to traverse brain tissue (Figure 2B). Multi-lead sEEG constructs may also constrain trajectories and increase the risk of brain shift (although thalamic leads are typically placed first). Within subgroups, ANT showed the greatest variability, likely due to anatomical heterogeneity,^46–48^ sharper lead entry angles, smaller cross-sectional area relative to the angle of approach, and differences in targeting methods—reliance on direct visualization of the mammillothalamic tract for ANT vs atlas-based targeting combined with direct visualization by FGATIR (Fast Gray Matter Acquisition T1 Inversion Recovery) and MP2RAGE (Magnetization-Prepared 2 Rapid Acquisition Gradient Echo) sequences, as available, for CM and PUL.

Accuracy improved modestly with increasing surgical experience and adoption of a robotic platform. A numerically higher degree of variability was observed in cases performed with the frameless stereotactic articulating arm vs. robot, with higher number of outliers (5/54 vs 1/84), suggesting a trend towards greater precision with robot, although this did not have statistical significance. Notably, accuracy did not differ significantly between pediatric and adult patients, or between single- and multi-lead thalamic implantations. Our sEEG accuracy results are likely similar to a prior study^49^ that reported a median target point error of 1.7 mm (note the difference in methodology; that study calculated Euclidian distance between planned target coordinates and the implanted lead, while here we measured distance between the lead and the full atlas volume of the targeted nucleus).

Distinct thalamocortical connectivity profiles were observed between the ANT, CM, and PUL thalamic nuclei. ANT was robustly connected to the prefrontal and mesial temporal regions; CM to peri-rolandic motor/sensory and the supplemental motor areas; and PUL to temporal and parieto-occipital regions. While ANT, CM, and PUL generally had distinct network connectivity patterns, some overlap was observed. For example, ANT and CM showed strong connectivity to the superior frontal gyrus, with a rostral (favoring ANT) to caudal (favoring CM) gradient. ANT also showed stronger connectivity than CM to rostral middle frontal and lateral orbitofrontal cortices. Similarly, CM and PUL shared overlapping projections to the parietal region, with CM parietal connectivity being largely restricted to the postcentral gyrus. The relative strength and hierarchy of thalamocortical projections between the sEEG and DBS cohorts were highly conserved for all subgroups (Figure 5), indicating that thalamic sEEG sampling is generally representative of DBS.

Some limitations exist in this single center study. Accuracy measurements used pragmatic electrode-to-atlas proximity metrics, rather than planned trajectory comparisons, due to limited surgical planning data. Normative structural connectome data were used as patient-specific connectivity data were not consistently available. Structural connectivity profiles do not indicate the directionality of pathways, which is critical when considering information flow through networks and the impact of neuromodulation. For instance, studies have shown non-reciprocal thalamocortical effective connectivity (e.g. strong hippocampus to pulvinar, but sparse pulvinar to hippocampus projections).^13,14,50^ Lastly, patients with severe structural brain abnormalities were excluded due to unreliable patient-to-template space normalization.

In conclusion, thalamic sEEG demonstrates a favorable safety profile, excellent target accuracy, and distinct target-specific thalamocortical connectivity across ANT, CM, and PUL in adult and pediatric patients. The higher rate of symptomatic surgical complications with thalamic DBS likely reflects instrumentation differences—particularly the larger cross-sectional area of DBS guide cannula. These findings provide a foundation for hypothesis-driven thalamic sampling during sEEG, while underscoring that intracranial electrode placement entails inherent risk and warrants individualized risk-benefit evaluation.

## Data Availability

Data will be made available upon reasonable request.

## Acknowledgements

None.

## Author Contributions

S. Park: study concept or design; major role in the acquisition of data; analysis or interpretation of data; drafting/revision of the manuscript for content. G. Ortiz-Guerrero: major role in the acquisition of data; drafting/revision of the manuscript for content. F. Permezel: analysis or interpretation of data; drafting/revision of the manuscript for content. H. Dabaja: major role in the acquisition of data; drafting/revision of the manuscript for content. G. Osman: drafting/revision of the manuscript for content. B. Burkett: drafting/revision of the manuscript for content. S. Messina: drafting/revision of the manuscript for content. K. Starnes: drafting/revision of the manuscript for content. B.N. Lundstrom: drafting/revision of the manuscript for content. L.C. Wong-Kisiel: drafting/revision of the manuscript for content. D. Burkholder: drafting/revision of the manuscript for content. D. Hermes: drafting/revision of the manuscript for content. B.H. Brinkmann: drafting/revision of the manuscript for content. G. Worrell: drafting/revision of the manuscript for content. W.R. Marsh: drafting/revision of the manuscript for content. K.J. Miller: drafting/revision of the manuscript for content. J.J. Van Gompel: drafting/revision of the manuscript for content. N.M. Gregg: study concept or design; analysis or interpretation of data; drafting/revision of the manuscript for content.

## Glossary

AAL3: Automated Anatomical Labeling atlas 3
ANT: anterior nucleus of thalamus
CM: centromedian nucleus
COM: center of mass
DBS: deep brain stimulation
FDR: false discovery rate
FGATIR: Fast Gray Matter Acquisition T1 Inversion Recovery
MNI: Montreal Neurological Institute
MP2RAGE: Magnetization-Prepared 2 Rapid Acquisition Gradient Echo
PUL: pulvinar nucleus
RNS: responsive neurostimulation
ROI: region of interest
sEEG: Stereoelectroencephalography
VTA: volume of tissue activated.

