## Supplementary Materials for "Thalamic stereoelectroencephalography: safety, accuracy, and thalamocortical connectivity"

**Subgroup analysis – comparison of surgical techniques**

The accuracy of sEEG implantation techniques were compared (Figure 4). Median proximity values for the stereotactic articulating arm, 3D printed stereotactic guide, or robotic system were 0.34 mm [IQR=0.23-1.04] (*n*=54), 0.32 mm [IQR=0.22-1.20] (*n*=22), 0.28 mm [IQR=0.19-0.41] (*n*=84), respectively (*p*=0.09). Pairwise Wilcoxon rank-sum test with Benjamini Hochberg false discovery rate (FDR) correction did not show a significant difference between articulating arm and robot (*p_FDR_*=0.11), articulating arm vs 3D printed stereotactic guide (*p_FDR_*=0.82) or robotic system vs 3D printed stereotactic guide (*p_FDR_*=0.34). There were 5 outliers in the articulating arm subgroup (9.3%), and 1 outlier each in the 3D printed stereotactic guide subgroup (4.6%) and robotic system subgroup (1.2%). Pairwise Fisher test showed a numerically higher outlier rate for articulating arm vs. robotic system (*p*=0.034, OR = 8.47, 95%, CI: 0.96-74.62), which was not significant after FDR correction (*p_FDR_*=0.10). There were no significant differences between articulating arm vs 3D printed stereotactic guide (*p_FDR_*=0.67) or robotic system vs. 3D printed system (*p_FDR_*=0.56).

**Subgroup analysis – comparison of pediatric and adult sEEG cases**

107 leads (66.9%) were implanted in 78 adults (age ≥18 years), and 53 leads (33.1%) were implanted in 31 pediatric patients. Median proximity was 0.29 mm [IQR=0.21-0.55] and 0.26 mm [IQR=0.20-0.42], respectively, (*p*=0.065).

**Subgroup analysis – single target vs multitarget sEEG cases**

sEEG cases were stratified by the number of thalamic targets sampled. Number of thalamic targets in each case ranged from 1-4 leads. Statistics were calculated at the electrode contact level (again, using the closest single contact to target per lead). 71 leads (44.4%) were implanted in single target cases; 62 leads (38.7%) for 2-target; and 27 leads (16.9%) for 4-target cases. For single target cases, median proximity of the electrodes was 0.33 mm [IQR=0.21-1.17]. For 2- and 4-target cases, median proximity of the electrodes was 0.30 mm [IQR=0.20-0.78] and 0.28 mm [IQR=0.20-0.38], respectively. Comparison of the proximity in single target and multitarget cases did not show a significant difference (*p*=0.26).

**Regression analysis – sEEG implant year**

Univariate regression analysis showed a statistically significant, but small improvement of accuracy over time (slope=-0.162 mm/year, *p*=0.024, *R^2^*=0.038) (Figure S3). Target-specific regression in ANT and CM subgroup did not show statistical significance (-0.185 mm/year, *p*=0.106 for ANT; -0.057 mm/year, *p*=0.811 for CM). PUL subgroup showed a significant but miniscule increase in proximity over time (0.026 mm/year, *p*=0.035).

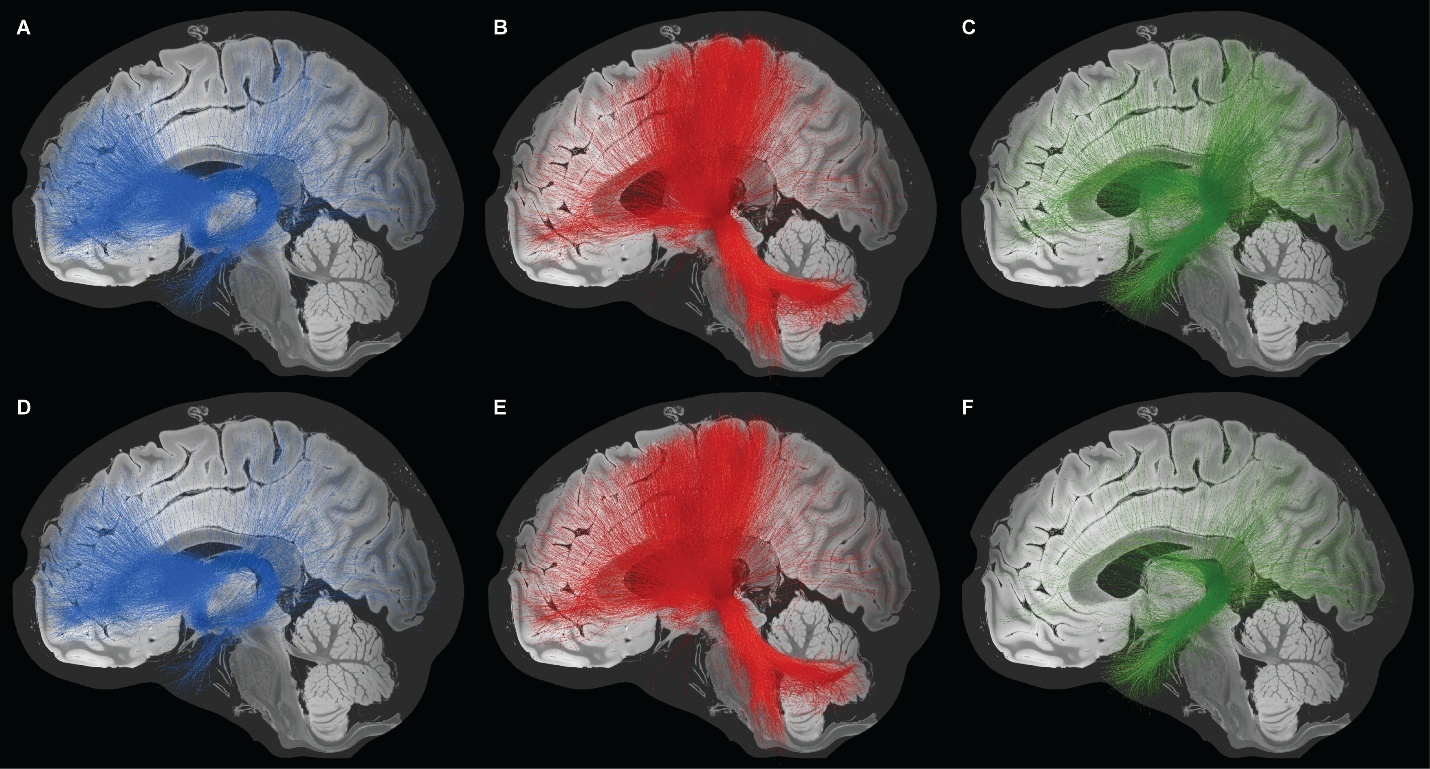

**Figure S1. Visualization of thalamocortical structural connectivity in sEEG (A, C, E) and DBS (B, D, F) cohort.** Group-level fiber filtering for each thalamic target and lead type, visualized against 7 Tesla MRI ex vivo human brain template.

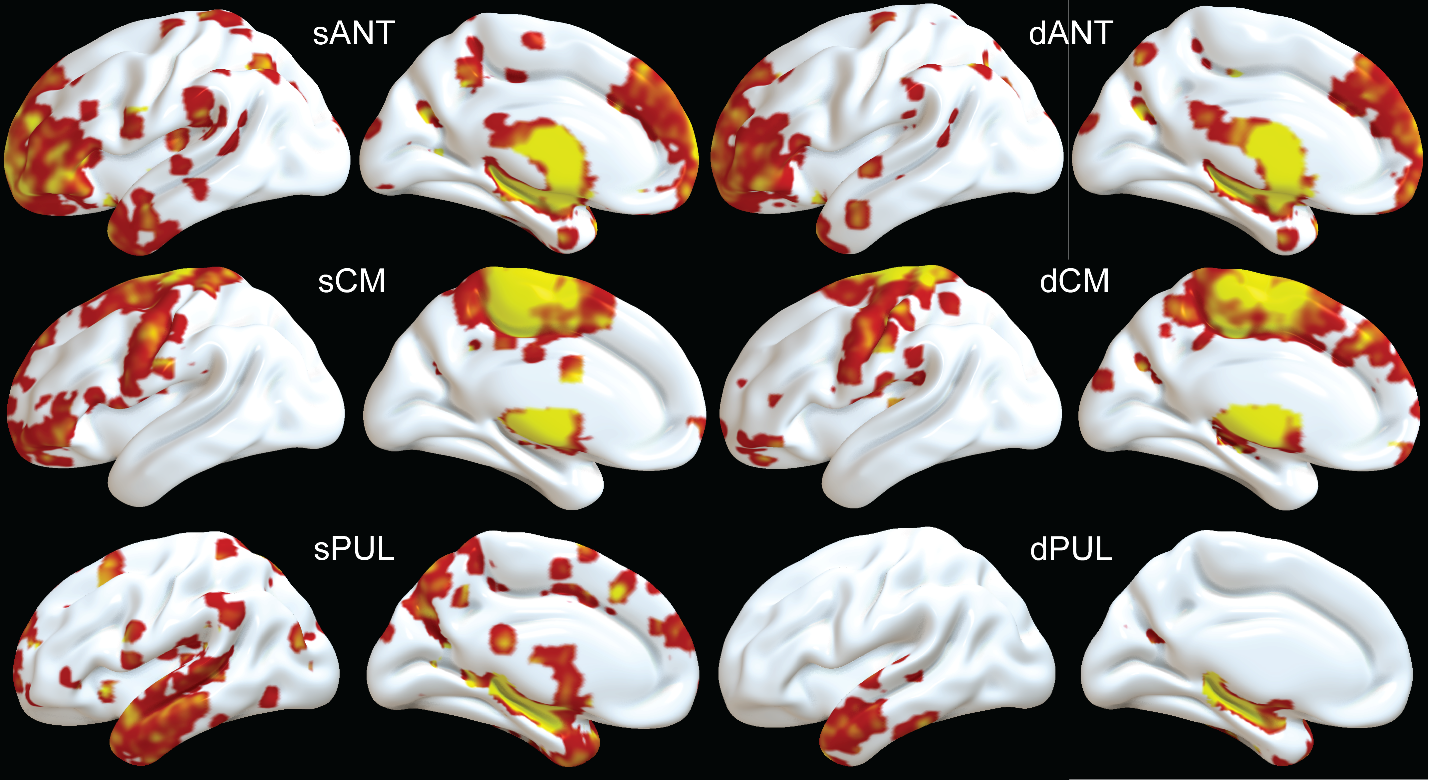

**Figure S2. Thalamocortical structural connectivity.** Group level thalamocortical structural connectivity was visualized on 3D cortical rendering using Surf Ice surface rendering tool (<https://www.nitrc.org/projects/surfice>). sANT, dANT: ANT subgroup in sEEG and DBS cohorts. sCM, dCM: CM subgroup in sEEG and DBS cohorts. sPUL, dPUL in sEEG and DBS cohorts.

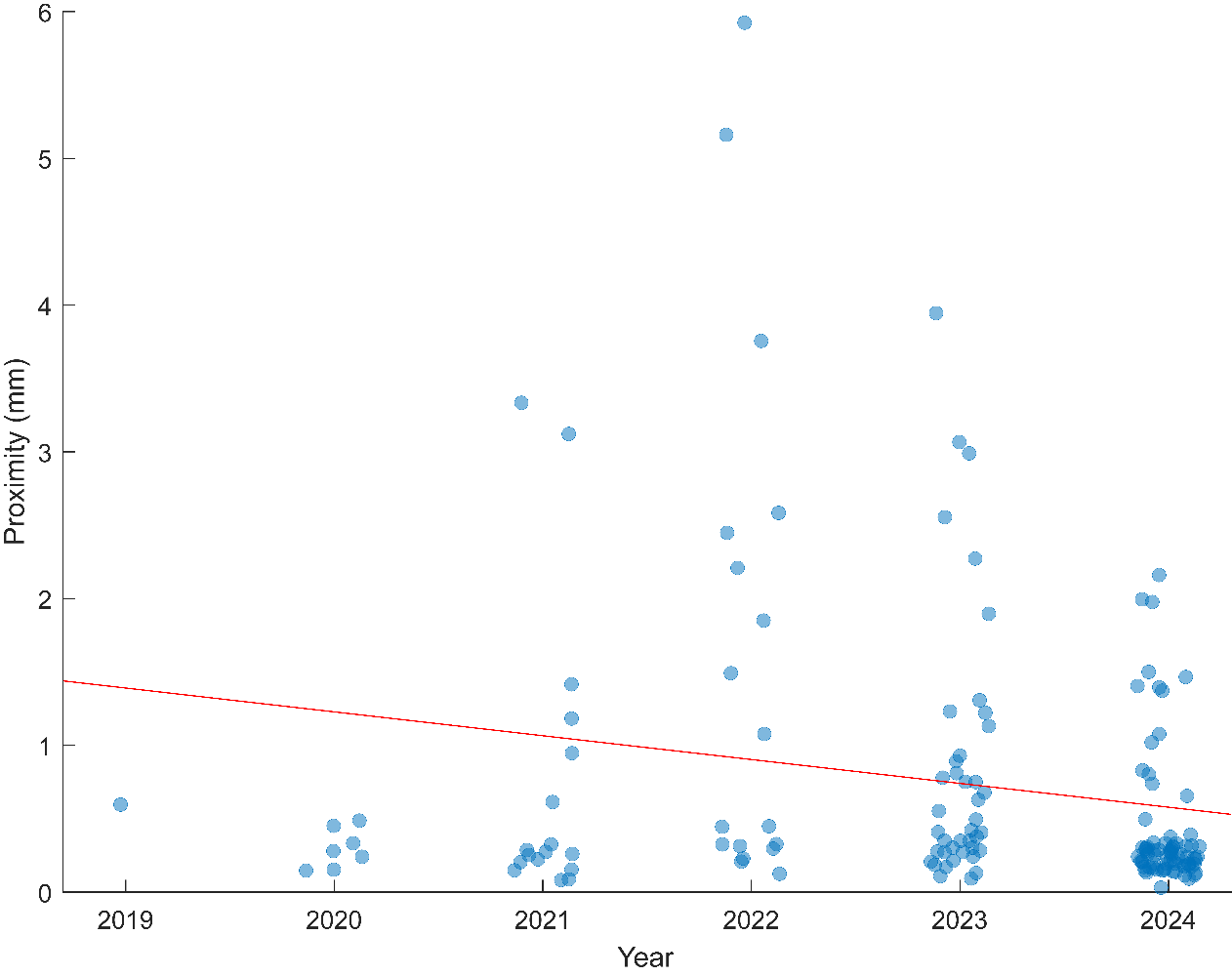

**Figure S3. Regression plot.** There was a small improvement of proximity (slope=-0.162 mm/year, *p*=0.024, *R^2^*=0.038).

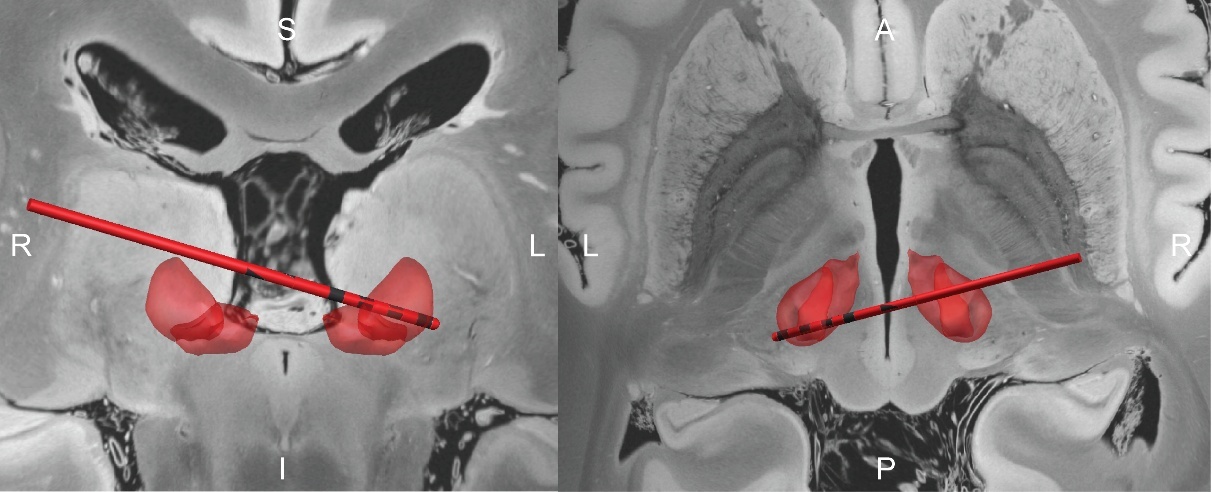

**Figure S4. Cross-hemispheric CM-DBS electrode.** One subject had a CM lead with cross-hemisphere trajectory due to multiple prior brain surgeries and lack of ipsilateral bone anchor, withheld from visualization in Figure 1 to avoid confusion.

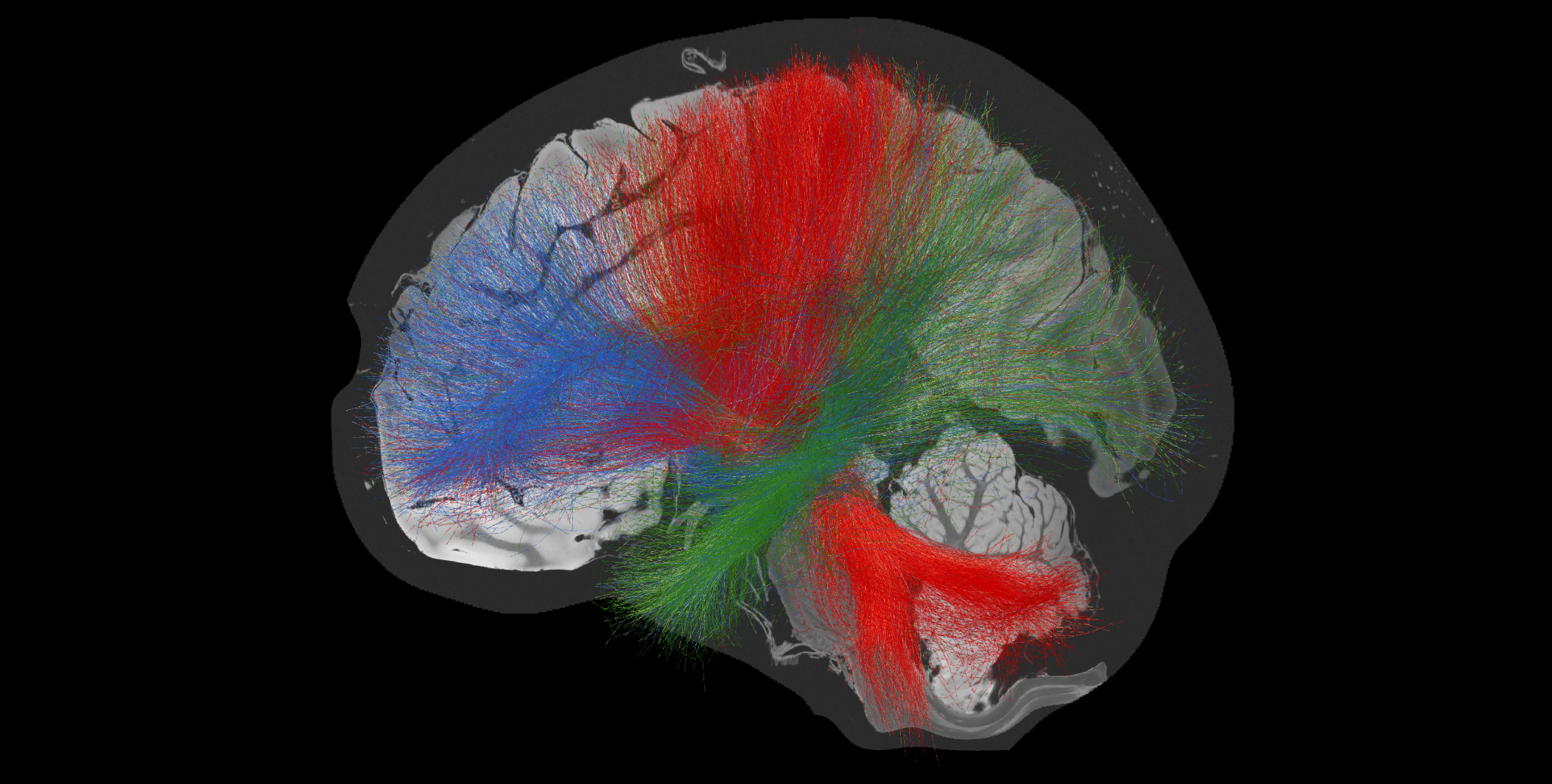

**Figure S5.** **Sagittal view of thalamocortical connectivity from ANT (blue), CM (red), PUL (green) in sEEG cohort.**
